# Dataset of COVID-19 outbreak and potential predictive features in the USA

**DOI:** 10.1101/2020.10.16.20214098

**Authors:** Arezoo Haratian, Hadi Fazelinia, Zeinab Maleki, Pouria Ramazi, Hao Wang, Mark A. Lewis, Russell Greiner, David Wishart

## Abstract

This dataset provides information related to the outbreak of COVID-19 disease in the United States, including data from each of 3142 US counties from the beginning of the outbreak (January 2020) until September 2020. This data is collected from many public online databases and includes the daily number of COVID-19 confirmed cases and deaths, as well as 33 features that may be relevant to the pandemic dynamics: demographic, geographic, climatic, traffic, public-health, social-distancing-policy adherence, and political characteristics of each county. We anticipate many researchers will use this dataset to train models that can predict the spread of COVID-19 and to identify the key driving factors.

## Background & Summary

With the current dramatic international COVID-19 epidemic, understanding the pandemic dynamics and improving preventive policies are crucial. An effective way to prevent the progression of the outbreak in the affected regions is to identify and if possible, control the factors influencing the spread of the disease in each region. However, the many factors that play a role in the spread of COVID-19 make it challenging to forecast, and hence plan for, the disease spread. Therefore, to examine the impact of potentially influential factors on the disease spread in the United States, we have collected a dataset containing, in each county and for each day since the beginning of the outbreak, the number of confirmed COVID-19 cases and deaths as well as 33 factors that may be relevant to the pandemic dynamics. In addition to the raw dataset, we have prepared a processed version of the dataset, where the missing values are imputed and the abnormal values, e.g., negative counting values, are fixed.

We anticipate this dataset will be useful for understanding, modeling, and predicting the COVID-19 pandemic dynamics in the United States at the “county” spatial resolution. This information may enable researchers and governments to gain a better understanding of the COVID-19 pandemic dynamics, which may help inform appropriate preventive policies. The dataset also can provide insights into the wide variety of potential factors affecting the spread of COVID-19. We anticipate its wide range of daily features, recorded over many months for a large number of counties, will be sufficient to estimate the parameter values of mechanistic models, such as the Susceptible-Infected-Recovered (SIR) type models^1^, as well as effective machine-learning models^2^.

## Methods

We rely on authoritative government and academic sources to collect the data, here is to provide the data for each of the features at the county level. For each of the 3142 counties in the US, for each day from the beginning of the diseaseouttrea in the country, January 22, 2020, until September 15, 2020, the dataset provides the number of confirmed COVID-19 cases and deaths as well as 33 other demographic, geographic, climatic, epidemiological and sociological features that potentially influence the spread and effects of the disease. These features include both fixed and temporal characteristics. Fixed (time-invariant) features generally represent a county’s geographic, demographic, and public health information. distancinv -adherence to social, actors eatures consist o climate)garyinv-eme(Temporal and a numter o tests per ormed in each, girus pressure rom neivhtorinv counees, policies state(Online-only Table 1).processinv o each o the gariatles are -The colleceon and pre .described below

**Online-only Table 1.** Description of the features.

| # | Variable Name | Description | Percentage of values available in the dataset | Type | Finest spatial scale |
| --- | --- | --- | --- | --- | --- |
| Target variables |  |  |  |  |  |
| (1) | COVID-19 confirmed cases | Number of daily COVID-19 confirmed cases | 100% | real | county |
| (2) | COVID-19 deaths | Number of daily COVID-19 deaths | 100% | real | county |
| Fixed features |  |  |  |  |  |
| (3) | Total population | Total population | 100% | real | county |
| (4) | Population density | Population per square mile | 100% | real | county |
| (5) | Female ratio | Total number of females divided by the total population | 100% | real | county |
| (6) | Age distribution | Percentage of residents in the age groups: 0-4, 5-9, 10-14, 15-19, 20-24, 25-29, 30-34, 35-39, 40-44, 45-49, 50-54, 55-59, 60-64, 65-69, 70-74, 75-79, 80-84, 85 and older | 100% | real vector (18 values, that add up to 1) | county |
| (7) | Education level distribution | Percentage of residents with different levels of education: 'less than high school diploma', 'high school diploma', 'some college or associate's degree' | 100% | real vector (4 values, that sum to 1) | county |
| (8) | Median household income | - | 100% | real | county |
| (9) | GDP per capita | Gross Domestic Product per capita (economic output divided by the population) | 100% | real | county |
| (10) | Area | Area in square miles | 100% | real | county |
| (11) | Latitude | Latitude of the county center | 100% | real | county |
| (12) | Longitude | Longitude of the county center | 100% | real | county |
| (13) | Housing density | Number of housing units per square mile (Including houses, apartments/flats, mobile homes, and other housing units) | 100% | real | county |
| (14) | Academic population ratio | Total number of residents who are currently university and college students or staff, divided by the total population | 100% | real | county |
| (15) | Immigrant students ratio | Total number of students who study in this county but are residents of the other states, divided by<br>the total county<br>population | 100% | real | county |
| (16) | Hospital bed<br>ratio | Number of Hospital beds<br>divided by the total<br>population | 100% | real | county |
| (17) | intensive care<br>unit (ICU) bed<br>ratio | Number of ICU beds<br>divided by the total<br>population | 77% | real | county |
| (18) | Ventilator<br>capacity ratio | Number of ventilators<br>divided by the total<br>population | 77% | real | county |
| (19) | Percent of<br>smokers | Percentage of adult<br>smokers | 100% | real | county |
| (20) | Percent of<br>diabetes | Percentage of diabetic<br>adults | 100% | real | county |
| (21) | Religious<br>congregation<br>ratio | Number of active<br>members of Religious<br>congregations divided by<br>the total population | 99% | real | county |
| (22) | Number of meat<br>plants | Number of meat<br>processing plants | 100% | discrete | county |
| (23) | Airport distance | Distance to the nearest<br>international airport with<br>average daily passenger<br>load more than ten | 100% | real | county |
| (24) | Passenger load<br>ratio | Average daily passenger<br>load of that nearest<br>international airport<br>divided by the total<br>population | 100% | real | county |
| (25) | Percent of<br>insured<br>residents | Percentage of health<br>insured residents | 99% | real | county |
| (26) | Deaths per<br>100,000<br>residents | Number of deaths per<br>100,000 residents | 76% | real | county |
| (27) | Political party | The political party of the<br>governor of each state (0<br>for Republican and 1 for<br>Democratic) | 100% | discrete | state |
| Temporal features |  |  |  |  |  |
| (28) | Precipitation | Daily precipitation | 65% | real | county |
| (29) | Temperature | Daily average temperature | 52% | real | county |
| (30) | Daily state test | Number of total COVID-19 tests performed at each day in the state of the county (including antibody, antigen, and PCR tests) | 82% | integer | state |
| (31) | Social distancing travel distance grade | Percent change in average distance traveled compared to pre-COVID-19-period (range from A to F)<br>A: >70% decrease<br>B: 55-70% decrease<br>C: 40-55% decrease<br>D: 25-40% decrease<br>F: <25% decrease or increase | 83% | nominal | county |
| (32) | Social distancing visitation grade | Percent change in non-essential visitation compared to pre-COVID-19 period (range from A to F)<br>A: >70% decrease<br>B: 65-70% decrease<br>C: 60-65% decrease<br>D: 55-60% decrease<br>F: <55% decrease or increase | 56% | nominal | county |
| (33) | Social distancing encounters grade | Percent change in human encounters compared to pre-COVID-19 period (range from A to F)<br>A: >94% decrease<br>B: 82%-94% decrease<br>C: 74%-82% decrease<br>D: 40%-74% decrease<br>F: <40% decrease or increase | 83% | nominal | county |
| (34) | Social distancing total grade | Average numerical score of the previous three social distancing factors | 83% | nominal | county |
| (35) | Virus pressure | A measure for virus transmission from neighboring counties, defined as the weighted average of the number of confirmed cases in the adjacent counties (ie, that share a border with this county) | 100% | real | county |

### Target Variables

We obtained the number of **COVID-19 confirmed cases** and **deaths** from the *USAFacts* website^3^, which is sourced from the *US Centers For Disease Control and Prevention (CDC)*^4^. This data source records the cumulative number of confirmed COVID-19 cases and the cumulative number of confirmed COVID-19 deaths for each county and day from the first case report on January 22, 2020. We obtained the new cases (and deaths) per day by subtracting the total of each day from the previous. This resulted in negative values for some counties on some days. After contacting the data source, we learned that the government agencies update their reported accumulative numbers to be lower than the previous day if they obtain a more accurate count by re-examining their medical records. Thus, the negative values generated in the daily confirmed cases and deaths data are related to the additional counts that were incorrectly reported. In the processed dataset, we used the following method to solve this problem. We set the entry of each day with a negative value to zero and add this negative value to the previous day’s entry. Then if the result of the sum is negative, we repeat this step until one day’s entry sums up to a positive value. So far, the repetition was never required as the resulting sums were always non-negative.

### Fixed Features

#### Demographic Features

We obtained most of the demographic data --The **total population, age** and **gender distribution**, number of housing units, and county **area** -- from the *US Census Bureau* websites; here we report 2018 values^5,6^. We calculated **population** and **housing density** by (respectively) dividing the total population and number of housing units in each county by the area of that county.

To calculate the **academic population ratio**, we first collected the total enrollment of university and college students and academic staff that each county reported in the fall of 2018 using data from the *National Center of Education Statistics (NCES)*^7^. This was divided by the population of that county in 2018 to calculate the ratio.

**Education level distribution** is the percentage of educated people at the three education legels ‘less than hivh school diploma’, ‘hivh school diploma’, and ‘some colleve or associate’s devree’, dhich de dodnloaded rom the *United States Department of Agriculture (USDA)*^8^. (Note this is 3 real values, each between 0 and 1.)

Immigrant students are those who enrolled in the fall of 2018 in any institution in the county for the first time at the undergraduate level but reside in another state^7^. We derived the **immigrant student ratio** by dividing the total number of immigrant students by the total county population.

**Religious congregation ratio** is calculated by dividing the total number of active members of a county’s relivious congregations reported in 2010 by the total county population^9^.

We collected from CDC^10^ the number of **deaths per 100**,**000 residents** as the overall death rate in 2018 for each county, regardless of the cause of the death.

#### Health Facilities and Risk Factors Features

The information about a county’s health facilities is included in the data mostly on a per capita basis. We collected the number of **intensive care unit (ICU) beds** and **ventilator capacity** data from the *Tableau Public* website^11^ and the total number of **hospital beds** per 1000 individuals from the *Urban Institute* website^12^, which is sourced from the *American Hospital Association Annual Survey* Database^13^. Then we derived the per capita information atout these acilities usinv each county’s population. **Percent of smokers** and **Percent of diabetes** show the percentage of adult smokers and diabetic adults in the total population of the county, respectively. Our data source for smoker and diabetes ratios and **percent of insured residents** was the *County Health Rankings and Roadmaps* website^14^.

#### Geographic Features

**Airport distance** for each county shows the distance to the nearest international airport, ottained ty considerinv the “vreat circle distance” calculated throuvh the latitude and longitude of the airport and the county center. Only airports with more than ten daily passengers, on average, are considered (using data prior to COVID-19).

The passenger load for each county is the passenger load of the nearest international airport to that county, and if a county includes more than one international airport, the passenger load equals the total passenger load of these airports. These data were collected from the *United States Department of Transportation* website and other resources^15,16,17^. We derived the **passenger load ratio** by dividing the passenger load by the total population of the county.

The **number of meat plants** shows the number of meat and poultry processing plants in each county, collected from the *United States Department of Agriculture* website^18^.

#### Economic features and other features

Our economic features for each county include the **median household income** and **GDP per capita**, both taken from data reported in 2018. These data are obtained from the *Census Bureau* and the *United States Bureau of Economic Analysis* and *State Science and Technology Institute* websites^19,20,21^. The governing **political party** data was collected from *wikipedia*^22^.

### Temporal Features

#### Climate Features

Our source for climate data contains daily **precipitation** and daily maximum, minimum, and average values for **temperature** each day. We obtained these data from the *Daily Summaries* dataset^23^. Note that 35% of the daily precipitation values were missing, which we imputed using the *KNN imputer*^24^ on the dataset consisting of the daily precipitation values over the counties. We similarly imputed each of the other temporal features using the KNN imputer on that temporal feature. As with daily average temperature, 82% of the values were missing. If the corresponding min and max daily temperatures were reported, we would impute the mean by averaging those two values. We used the KNN imputer to impute the remaining 48% of the missing values for the instances that did not include any of the average, minimum, or maximum temperatures.

#### Social Distancing Features

Our social distancing data source, *Unacast*^25^, is based on mobile location data.

In collecting this data, the users consent to opt-in and can opt-out by filling out a form on the data source website. This data source contains four different metrics: **travel distance grade, visitation grade, encounters grade**, and an overall average score of these three metrics. Each of these grades determines the percentage of reduction in a measure of unnecessary social activities (e.g. traveling, human encounters, non-essential visitation) compared to the pre-COVID-19 period, which is translated into letter grades, as described in Online-only Table 1.

The **encounters grade** is based on the proximity of two devices within a circle of radius 50m for less than an hour, counted as one encounter. This grade shows the decrease in human encounter density (number of encounters per square km) compared to the baseline, where the baseline is defined as the national average encounter density during the four weeks before the COVID-19 outbreak (February 10th - March 8th). The reason why the baseline is defined over the whole nation is to assign lower grades to denser areas, even if they witness fewer encounters compared to the pre-COVID-19 period. Namely, dense areas have a high infection risk, even if they were denser in the past.

The **visitation grade** indicates the percent change in visits to non-essential venues compared to the pre-COVID-19 period. Non-essential venues include but are not limited to restaurants (multiple kinds), department and clothing stores, jewelers, consumer electronics stores, cinemas and theaters, office supply stores, spas and hair salons, gyms and fitness/recreation facilities, car dealerships, hotels, craft, toy and hobby shops. The average visitation for each day of the week prior to the COVID-19 outbreak (between February 10th to March 8th) is considered as the baseline. The percent change is calculated by comparing those baselines to visits on the corresponding days of the week post-outbreak (March 9th onwards).

The **travel distance grade** simply shows the percentage reduction in average distance traveled in each county for each day. The highest grade for this metric represents more than 70% reduction in average distance traveled. This threshold is selected based on the experience gained from Italy because they implemented some of the most strict social distancing policies, which resulted in a 70% to 80% reduction. Therefore, Unacast expects a maximum of 70% reduction in distance traveled under a total shot-down.

The data source began recording the social distancing data from February 24. We set the values for the previous days (January 22 to February 23) to the lowest grade for each social distancing feature, assuming that no social distancing policies were imposed by then. However, the **encounters grade, travel distance grade**, and **total grade**, each still had 17% missing values for the remaining dates (post 24 February) and the **visitation grade** had 44% missing values, all of which we imputed using the KNN imputer.

#### Other features

**Daily state tests** refer to the number of daily tests performed in each state. These numbers were obtained using statistics from multiple type COVID-19 tests including antibody, antigen, and PCR. This data was downloaded from the *COVID Tracking Project*^26^ and contains negative values for some states and days. The starting recording date for the number of performed tests varies from state to state. We considered the negative numbers as missing values and imputed them along with the unrecorded number of tests using the KNN imputation method.

The **virus pressure** at county *x*_*i*_, and day *t*, denoted by *V*(*x*_*i*_,*t*), is defined based on the number of COVID-19 cases in the neighboring counties:

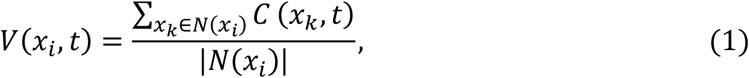

where *C*(*x*_*k*_,*t*)denotes the number of COVID-19 cases in county *x*_*k*_ at day *t*, and *N*(*x*_*i*_) is the set of the neighboring counties of county *x*_*i*_.

We removed from the processed dataset, those counties that missed any of the fixed features. For each of the temporal features, if a county had missing values for only some of the recorded dates, these missing values were imputed as explained above; however, if it missed the values in all of the recorded dates, the county was removed from the dataset. This resulted in the elimination of a total of 790 counties.

## Data Records

The version of the dataset at the time of submission, containing data from January 22 to September 16, has been archived in figshare^27^, and the latest version of the dataset is publicly available in our Github repository https://github.com/network-and-Data-Science-IUT/covid-19. We included 2 datasets: (i) the raw dataset with the negative and missing values, and (ii) the processed dataset where the counties with missing values are all imputed or removed from the dataset. Each row in the datasets corresponds to a specific county and date. Counties and their associated states are represented using their name and fips code^28^. Online-only Table 1 specifies the name, type, spatial scale, and description of each feature and also the percentage of their values available in the raw dataset. Note that virus pressure exists only in the processed dataset, and the visitation grade is removed from this dataset because it has a high volume of missing values. The size of the current raw and processed datasets are 277 and 218 MB. We plan to update the datasets until the end of 2021.

For illustration, Figure 1 plots the daily temporal features for the New York and Los Angeles counties over the peak days of the disease outbreak.

**Fig. 1:**
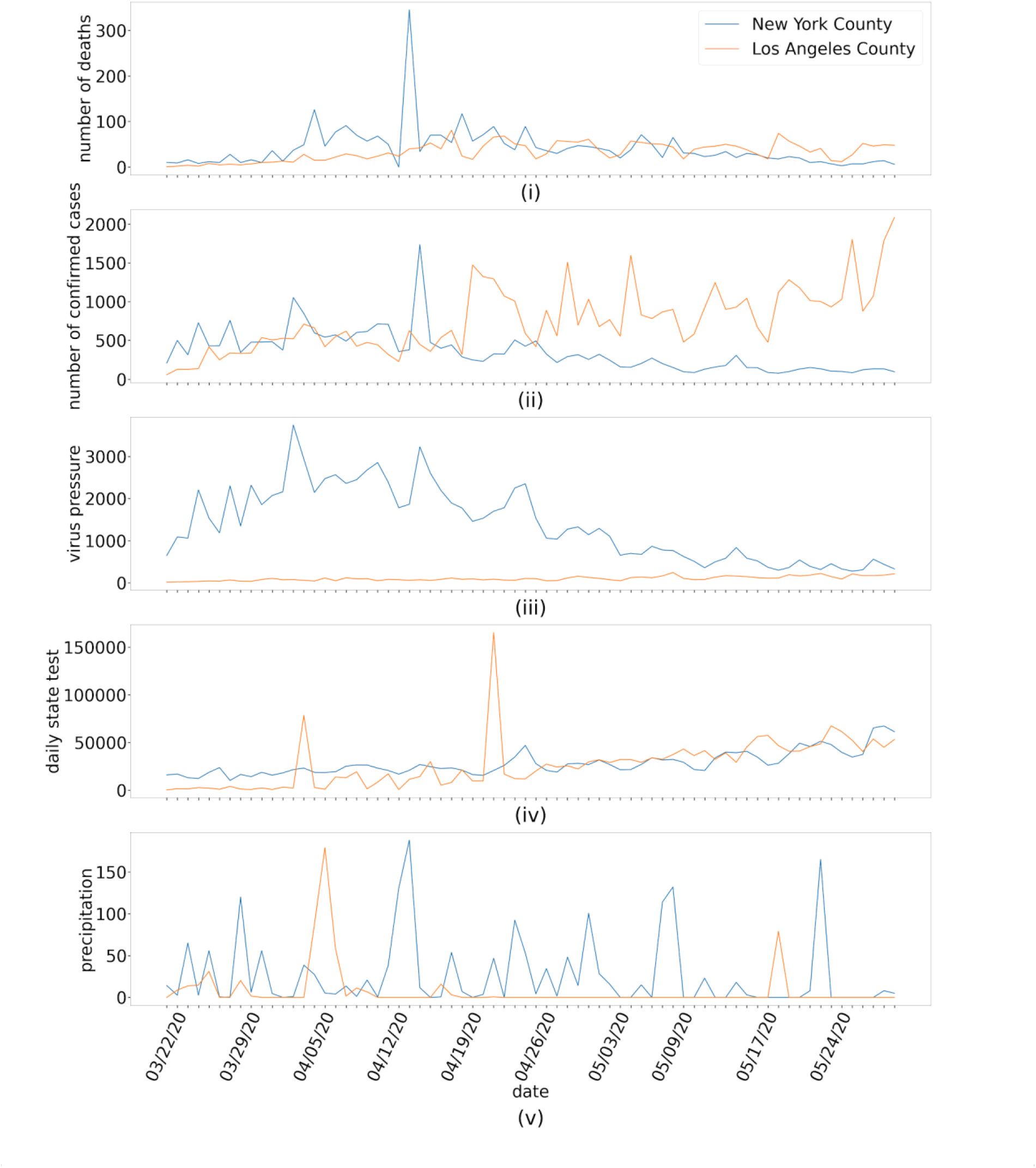
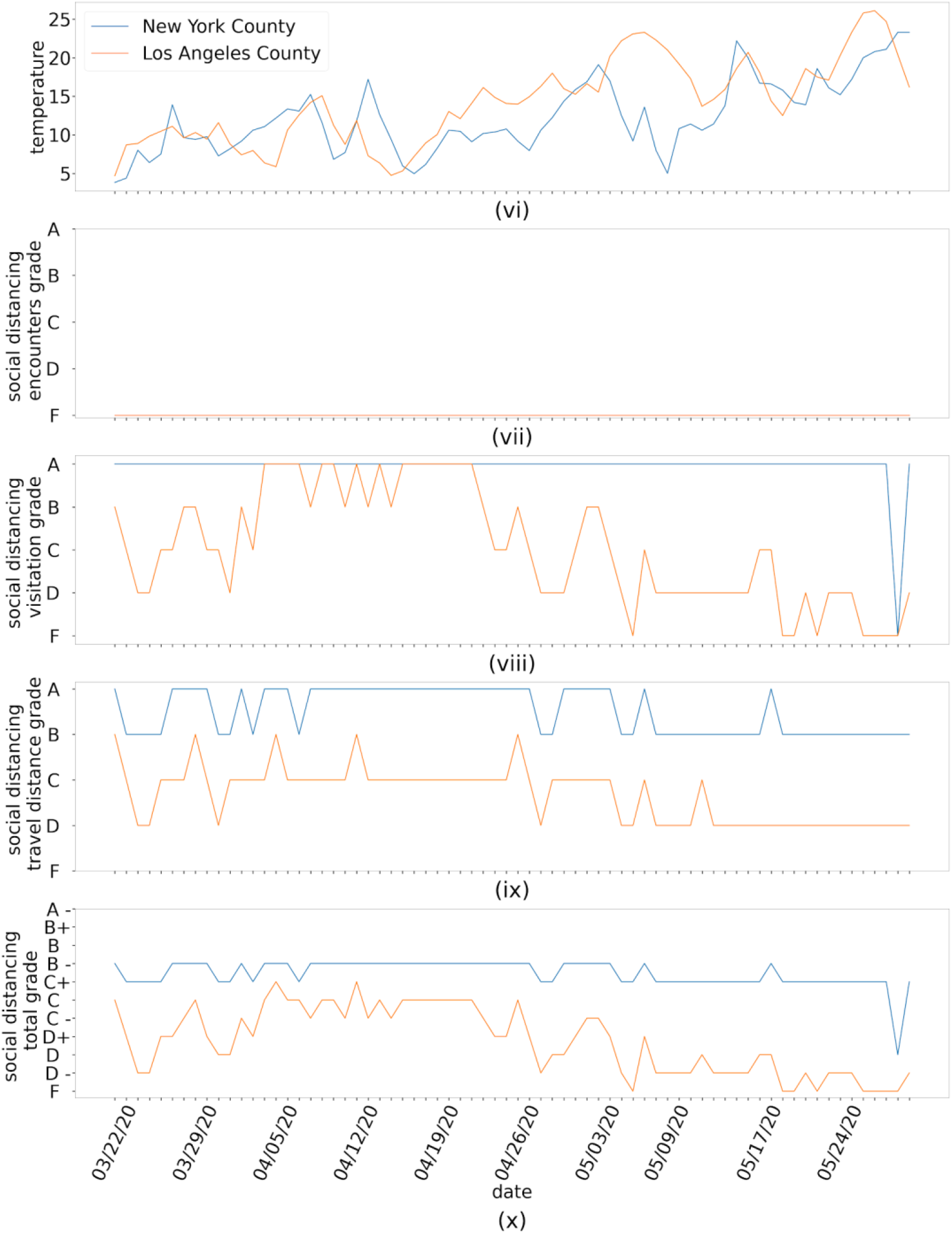
Evolution of the temporal features during the peak days of the COVID-19 outbreak (March 22, 2020, to May 30, 2020). (i) number of deaths, (ii) number of confirmed cases, (iii) virus pressure, (iv) daily state test, (v) precipitation, (vi) temperature, (vii) social distancing encounter grade, (viii) social distancing visitation grade, (ix) social distancing travel distance grade, (x) social distancing total grade.

## Technical Validation

We checked the compatibility of the number of COVID-19 confirmed cases and deaths with *worldometers* website^29^ reports. We checked the values of each collected feature for abnormal values (e.g., negative values for counting variables) using the *pandas* package^30^ for the *python* programming language. We then considered these values as missing values and imputed them using the imputation methods mentioned above. When adding a feature to the dataset, we used the pandas package to identify and remove duplicate feature records for the same county and date. In addition, by looking at the summary of each feature, including its min, max, and mean values, as well as randomly observing some of the values of that feature, we checked if the values belong to the logical range of that feature. For example, the raw data collected for temperature included values in the range [-500,500] for Celsius. After contacting the corresponding website^23^, it appeared that we had to divide by 10 to obtain the correct values in Celsius. None of the other features had this issue.

## Data Availability

The latest version of the dataset is publicly available in our Github repository https://github.com/network-and-Data-Science-IUT/covid-19

## Code Availability

Data collection and preparation are done using the python programming language. We used the *json* and *requests* packages^31,32^ to collect data and the *scikit-learn* package^33^ to impute missing values. The codes used to collect and prepare the datasets are available in our Github repository https://github.com/network-and-Data-Science-IUT/covid-19.

## Acknowledgments

This work was funded by Alberta Innovates, project number: RES0052027. We thank Dr. Ammar Hassanzadeh Keshteli for illustrative discussions.

## Author contributions

A.H. found relevant data sources and drafted the original manuscript. H.F. helped with finding the data sources, developed the code for automatic data collection, and contributed to the writing of the manuscript. P.R. and Z.M. managed and coordinated the production of the dataset and revised the manuscript. H.W., M.L., R.G., and D.W. revised the manuscript and supervised the study.

## References

1. Cooper, I., Mondal, A. & Antonopoulos, C.G. A SIR model assumption for the spread of COVID-19 in different communities. Chaos, Solitons & Fractals. 139, 110057 (2020).

2. Yadav, M., Perumal, M. & Srinivas, M. Analysis on novel coronavirus (COVID-19) using machine learning methods. Chaos, Solitons & Fractals. 139, 110050 (2020).

3. USA Facts, US Coronavirus Cases and Deaths. https://usafacts.org/visualizations/coronavirus-covid-19-spread-map/ (2020).

4. Centers For Disease Control and Prevention (CDC), CDC COVID Data Tracker. https://covid.cdc.gov/covid-data-tracker/?CDC_AA_refVal=https%3A%2F%2Fwww.cdc.gov%2Fcoronavirus%2F2019-ncov%2Fcases-updates%2Fcases-in-us.html#county-map (2020).

5. United States Census Bureau, Population and Housing Unit Estimates Tables. https://www.census.gov/programs-surveys/popest.html (2020).

6. United States Census Bureau, USA Counties: 2011.https://www.census.gov/library/publications/2011/compendia/usa-counties-2011.html#LND(2020).

7. National Center of Education Statistics (NCES). https://nces.ed.gov/ipeds/use-the-data (2020).

8. United States Department of Agriculture, Economic Research Service. https://www.ers.usda.gov/webdocs/DataFiles/48747/Education.xls?v=568.3 (2020).

9. Grammich, C. K. et al. U.S. Religion Census Religious Congregations and Membership Study, 2010 (County File). OSF https://doi.org/10.17605/OSF.IO/QUN29 (2018).

10. Centers for Disease Control and Prevention, Multiple Cause of Death, 1999-2018 Request.https://wonder.cdc.gov/controller/datarequest/D77 (2020).

11. Tableau Public, Definitive HealthCare COVID-19 Capacity Predictor. https://public.tableau.com/profile/todd.bellemare#!/vizhome/DefinitiveHCCOVID-19CapacityPredictor/DefinitiveHealthcareCOVID-19CapacityPredictor (2020).

12. Urban Institute. https://www.urban.org/policy-centers/health-policy-center/projects/understanding-hospital-bed-capacities-nationwide-amid-covid-19 (2020).

13. American Hospital Association Annual Survey 2018.https://www.ahadata.com/aha-annual-survey-database” https://www.ahadata.com/aha-annual-survey-database (2020).

14. County Health Rankings and Roadmaps. https://www.countyhealthrankings.org/app/ (2020).

15. Department of Transportation, International_Report_Passengers. https://data.transportation.gov/Aviation/International_Report_Passengers/xgub-n9bw (2020).

16. OpenFlight.org, Airport, airline and route data. https://openflights.org/data.html (2020).

17. Data.gov.https://catalog.data.gov/dataset/airports (2020).

18. United States Department of Agriculture, Food Safety and Inspection Service. https://origin-www.fsis.usda.gov/wps/wcm/connect/3e414e13-d601-427c-904a-319e2e84e487/MPI_Directory_by_Establishment_Name.xls?MOD=AJPERES (2020).

19. United States Bureau of Economic Analysis (BEA) https://www.bea.gov/data/gdp/gdp-county-metro-and-other-areas (2020).

20. State Science & Technology Institute (SSTI). https://ssti.org/blog/useful-stats-10-year-changes-real-gdp-county-and-industry-2009-2018 (2020).

21. United States Census Bureau, Small Area Income and Poverty Estimates (SAIPE) Program.https://www.census.gov/programs-surveys/saipe.html (2020).

22. List of United States governors. https://en.wikipedia.org/wiki/List_of_United_States_governors (2020).

23. National Climatic Data Center of National Oceanic and Atmospheric Administration.https://www.ncdc.noaa.gov/cdo-web/datatools/selectlocation (2020).

24. Troyanskaya, O. et al. Missing value estimation methods for DNA microarrays. Bioinformatics. 17.6, 520–525 (2001).

25. Unacast Company. https://www.unacast.com/covid19 (2020).

26. The COVID Tracking Project. https://covidtracking.com/ (2020).

27. Ramazi, P., Maleki, Z., Fazelinia, H., Haratian, A. USA covid-19 data. Figshare https://doi.org/10.6084/m9.figshare.12986069.v1 (2020).

28. FIPS county code. https://en.wikipedia.org/wiki/FIPS_county_code (2020).

29. worldometer. https://www.worldometers.info/coronavirus/country/us (2020).

30. McKinney, W. Data structures for statistical computing in python. Proceedings of the 9th Python in Science Conference. 445, 51–56 (2010).

31. json — JSON encoder and decoder. https://docs.python.org/3/library/json.html (2020).

32. Requests: HTTP for Humans. https://requests.readthedocs.io/en/master/ (2020).

33. Pedregosa, F. et al. Scikit-learn: machine learning in python. the Journal of machine Learning research. 12, 2825–2830 (2011).

